# IgM autoantibodies recognizing ACE2 are associated with severe COVID-19

**DOI:** 10.1101/2020.10.13.20211664

**Authors:** Livia Casciola-Rosen, David R. Thiemann, Felipe Andrade, Maria Isabel Trejo Zambrano, Jody E. Hooper, Elissa K. Leonard, Jamie B. Spangler, Andrea L. Cox, Carolyn E. Machamer, Lauren Sauer, Oliver Laeyendecker, Brian T. Garibaldi, Stuart C. Ray, Christopher A. Mecoli, Lisa Christopher-Stine, Laura Gutierrez-Alamillo, Qingyuan Yang, David Hines, William A. Clarke, Richard Rothman, Andrew Pekosz, Katherine J. Fenstermacher, Zitong Wang, Scott L. Zeger, Antony Rosen

**Author notes:** denotes corresponding authors; Livia Casciola-Rosen, Scott Zeger and Antony Rosen.

## Abstract

SARS-CoV-2 infection induces severe disease in a subpopulation of patients, but the underlying mechanisms remain unclear. We demonstrate robust IgM autoantibodies that recognize angiotensin converting enzyme-2 (ACE2) in 18/66 (27%) patients with severe COVID-19, which are rare (2/52; 3.8%) in hospitalized patients who are not ventilated. The antibodies do not undergo class-switching to IgG, suggesting a T-independent antibody response. Purified IgM from anti-ACE2 patients activates complement. Pathological analysis of lung obtained at autopsy shows endothelial cell staining for IgM in blood vessels in some patients. We propose that vascular endothelial ACE2 expression focuses the pathogenic effects of these autoantibodies on blood vessels, and contributes to the angiocentric pathology observed in some severe COVID-19 patients. These findings may have predictive and therapeutic implications.

**One-sentence summary:** ACE2 autoantibodies in severe COVID-19 have features of a T-independent immune response, and may mediate vascular damage.

---

COVID-19 is a global pandemic caused by the novel coronavirus SARS-CoV-2(*1*). It is a highly infectious pathogen, and continues to have a massive global impact since its recognition in Wuhan in late 2019, with more than 37 million confirmed infections, >1 million confirmed deaths and massive economic disruption across the world. While most infections appear to be self-limited, 15-20% of symptomatic individuals become hospitalized, and 5-10% require admission to ICUs(*2*),(*3*). Mortality rates of hospitalized patients in the US range between 13 and 28%. Growing evidence suggests that some of the severe COVID-19 clinical features represent damage induced by activation of the immune and inflammatory responses initiated by the virus(*4*),(*5*),(*6*). In addition to frequent acute respiratory distress syndrome (ARDS), there is also evidence of vasculopathy(*7*),(*8*), clotting(*9*),(*10*), and cardiovascular complications(*11*) whose mechanisms are presently unclear, but in which complement activation has been implicated(*12*),(*9*). The recent finding that low-dose dexamethasone has a beneficial effect on mortality in a subgroup of patients with severe COVID-19 requiring ventilation has suggested that uncontrolled inflammatory mechanisms might play an apical role in mediating disease severity in a subset of patients with this disease(*13*). Understanding these mechanisms is therefore a high priority, particularly if they might be rapidly addressed therapeutically with additional off-the-shelf approaches.

## IgM autoantibodies recognizing ACE2 are associated with severe disease in COVID-19

We were immediately drawn to ACE2, the host receptor for SARS-CoV-2 entry (*1*), as a potential autoantigen in COVID-19. SARS-CoV-2 spike (S) protein binds with higher affinity (5-20 fold higher) to ACE2 than the other coronaviruses which also bind to this host receptor (*14*). Furthermore, ACE2 expression is enhanced in lung (epithelial and endothelial cells) and heart (endothelial cells) (*15*), and hypomorphic ACE2 function has been implicated in adverse outcomes in models of ARDS (*16*). We therefore established assays to screen for IgM and IgG autoantibodies to ACE2, and applied these to a cohort of 66 hospitalized patients with COVID-19 that reached the 6 most severe WHO ordinal categories as their maximal severity (28 severe, 38 moderate). 8 patients were positive for ACE2 IgM autoantibodies. 7 of these were in the mechanically ventilated (WHO 6/7) or dead groups (WHO 8) (7/28; 25%), while only a single patient was positive among the 38 patients who were not ventilated (1/38; 2.6%; OR 12.3, 95% CI 1,875-141.9; p=0.0084; Fisher’s exact test; Supp.Fig. 1A). In order to increase sample size and define the stability and kinetics of these antibodies, we assembled additional patients in whom serum was available from multiple laboratory blood draws taken across their hospitalization. This added 52 COVID-19 patients for analysis (38 in WHO ordinal groups 6-8 [31 ventilated and 7 dead], and 14 patients in ordinal category 4). The frequencies of anti-ACE2 IgM in these patients were very similar to the initial group: 11/38 (28.9%) of the patients with severe COVID-19 were positive for anti-ACE2 IgM antibodies compared to 1/14 (7.1%) in the milder COVID-19 group (Supp. Fig. 1B). The combined frequency of anti-ACE2 IgM in severe COVID-19 was 18 of 66 patients (27.2%) compared to 2 of 52 patients with moderate COVID-19 (3.8%; p= 0.0009; OR 9.38, 95% CI 2.38-42.0; Fisher’s exact test; Fig. 1A). IgM levels were robust (Fig. 1A, center panel); all positives were confirmed and quantified by serial dilution (representative examples in Supp. Fig. 1B). Anti-ACE2 IgG were found in 12/66 (18%) patients with severe COVID-19 (WHO 6-8), and 6/52 (11.5%) patients with moderate disease (WHO 3-5; p=0.44, Fisher’s exact test). Only 4/18 (22%) severe patients with anti-ACE2 IgM antibodies were also IgG positive (Fig. 1A, right panel). ACE2 is therefore a prominent autoantibody target in patients with COVID-19, with IgM autoantibodies quite strikingly associated with severe disease.

**Figure 1:**
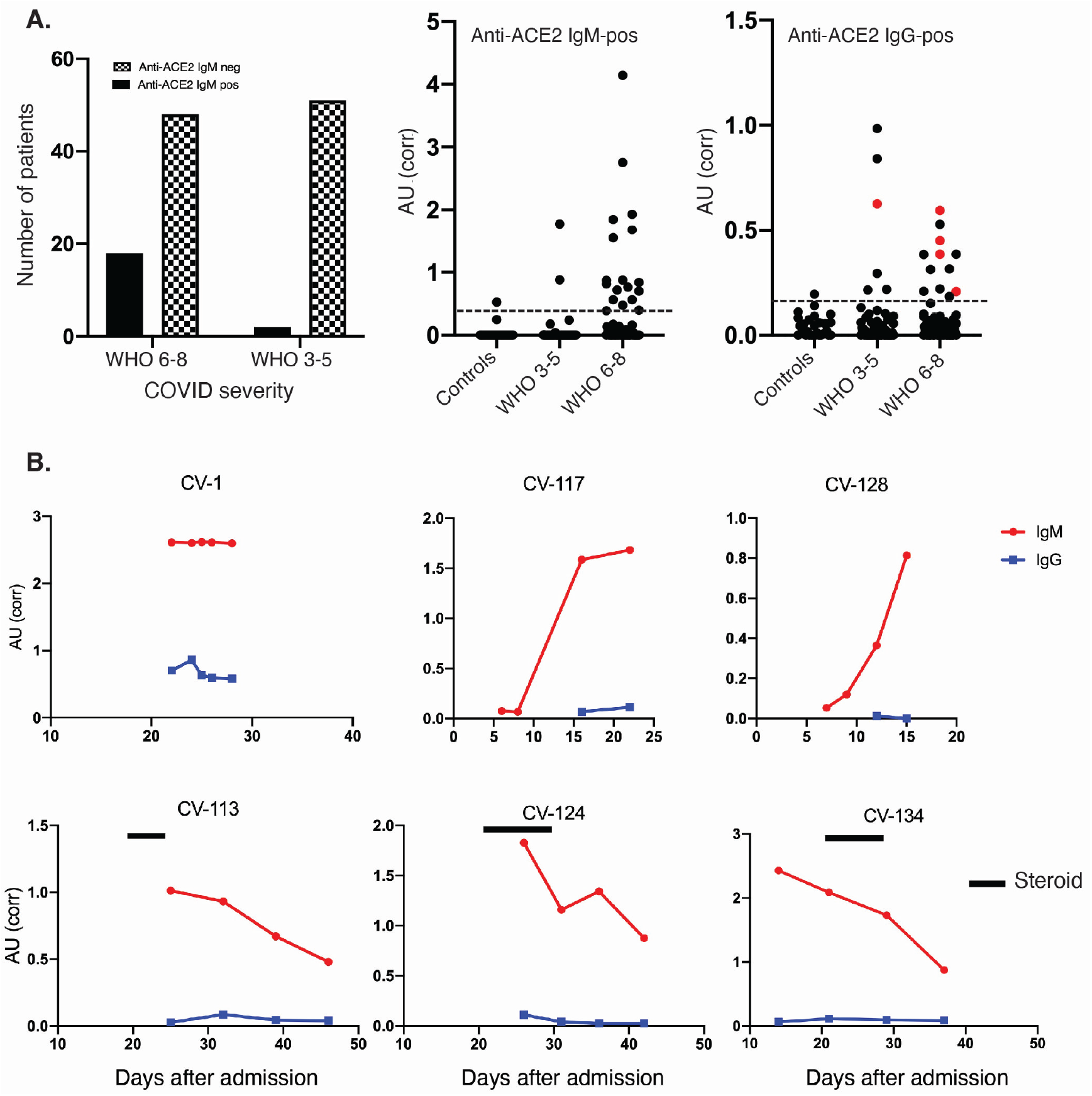
Anti-ACE2 IgM antibodies are found in patients with COVID-19. A: Antibodies were assayed by ELISA in the combined COVID cohort (N=118 patients). Left panel: the number of patients with and without anti-ACE2 IgM antibodies is shown grouped by disease severity. 27.2% of severe patients were anti-ACE2 positive compared to 3.8% with moderate COVID (p= 0.0009; Fisher’s exact test). In the center and right panels, data from anti-ACE2 IgM and IgG ELISA assays, respectively, is presented as corrected OD 450 absorbance units. This data was obtained on all the COVID patients presented in the left panel, as well as from 30 healthy controls. Red dots in the IgG panel denote IgG-positive samples that also have anti-ACE2 IgM antibodies. The horizontal line on each plot represents the cutoff for assigning a positive antibody status. B: Longitudinal analysis of anti-ACE2 IgM antibodies. For all those anti-ACE2 IgM-positive patients with multiple banked sera available (16/18), anti-ACE2 IgM and IgG antibodies were quantitated over time. Red and blue lines on each plot denote anti-ACE2 IgM and IgG antibodies, respectively. Solid black bars represent steroid treatment periods. Additional examples are shown in Suppl Fig 3.

Clinical features of the anti-ACE2 IgM-positive group are summarized in Fig 2 and Supp. Table 1. The mean age of the anti-ACE2 IgM-positive group was 61.5 years (N = 20, se = 9.7, S^2^ = P 93.6), compared to 59.0 (N = 98 se = 17.3, S^2^ = 298.8) years for IgM-negatives (t = 0.89, p = 0.37, unpaired t-test). 72% of anti-ACE2 IgM were present in females. While the proportion of anti-ACE2 was higher in females (13/38, 34%) than males (5/28, 17.8%) with severe COVID-19, this difference did not reach statistical significance in this sample (p=0.17; Fisher’s exact test). The mean BMI of IgM-positive patients was 35.4 (N = 16, se = 10.7, S^2^ = 115.2), compared to 30.4 (N = 81, se = 8.1, S^2^ = 65.2) in IgM-negative patients (t = 1.74, p = 0.10, unpaired t-test). Interestingly, the anti-ACE2-positive group had statistically significantly higher average temperatures over the first 10 days of hospitalization than the IgM-negative group (IgM-positive: mean = 37.5, S^2^ = 0.65, N = 783 on M = 20 unique patients, IgM-negative: mean= 37.0, S^2^P P =0.56, N = 3137 on M = 97 unique patients; chisq = 22.72, p = 0.0001 from linear mixed-effects model Wald test with 4 degrees of freedom (see statistical methods); Fig. 2D). The results did not qualitatively change when we restricted the analysis to the severe IgM-positive patients above and compared them to all severe COVID-19 patients from the CROWN Registry for whom IgM status was unknown (IgM-positive: mean = 37.53, S^2^ = 0.64, N = 721 on M = 18 unique patients, IgM-unknown: mean = 37.11, S^2^ =0.59, N =14827 on M = 473 unique patients; chisq = 19.98, p = 0.0005 from linear mixed-effects model Wald test with 4 degrees of freedom (see statistical methods), (Supp Fig. 2). Population average CRP levels were also different in the 2 groups in the first 10 days after admission, with the population average peaking at ∼d4-d6 after admission at 20mg/dL in the IgM-positive group, compared to 7.4mg/dL for the IgM-negative group (IgM-positive: mean = 16.96, S^2^=104.55, N = 95 on M = 18 unique patients, IgM-negative: mean = 13.52, S^2^ = 151.58, N = 413 on M = 90 unique patients; chisq = 11.19, p = 0.02, from linear mixed-effects model Wald test with 4 degrees of freedom (see statistical methods), Fig. 2E). Various infectious and autoimmune disease controls were also tested for anti-ACE2 IgM (Fig. 2F). Anti-ACE2 IgM autoantibodies were not observed in 30 patients with acute influenza infection (including 11 patients evaluated in the ED and discharged to outpatient care, and 19 hospitalized patients requiring oxygen therapy or assisted ventilation (5)), 25 patients with systemic lupus erythematosus (SLE), 13 with scleroderma, and 15 with autoimmune necrotizing myopathy. Interestingly, we did find ACE2 autoantibodies in an index patient with a rare acute dermatopulmonary syndrome associated with autoantibodies to MDA5 (*17*) from who we had collected serum previously, and which appears to phenocopy several of the features of severe COVID-19 (see case report in Methods; additional studies on similar patients are underway). This specificity of anti-ACE2 IgM for severe COVID-19 or a close phenocopy is striking.

**Figure 2:**
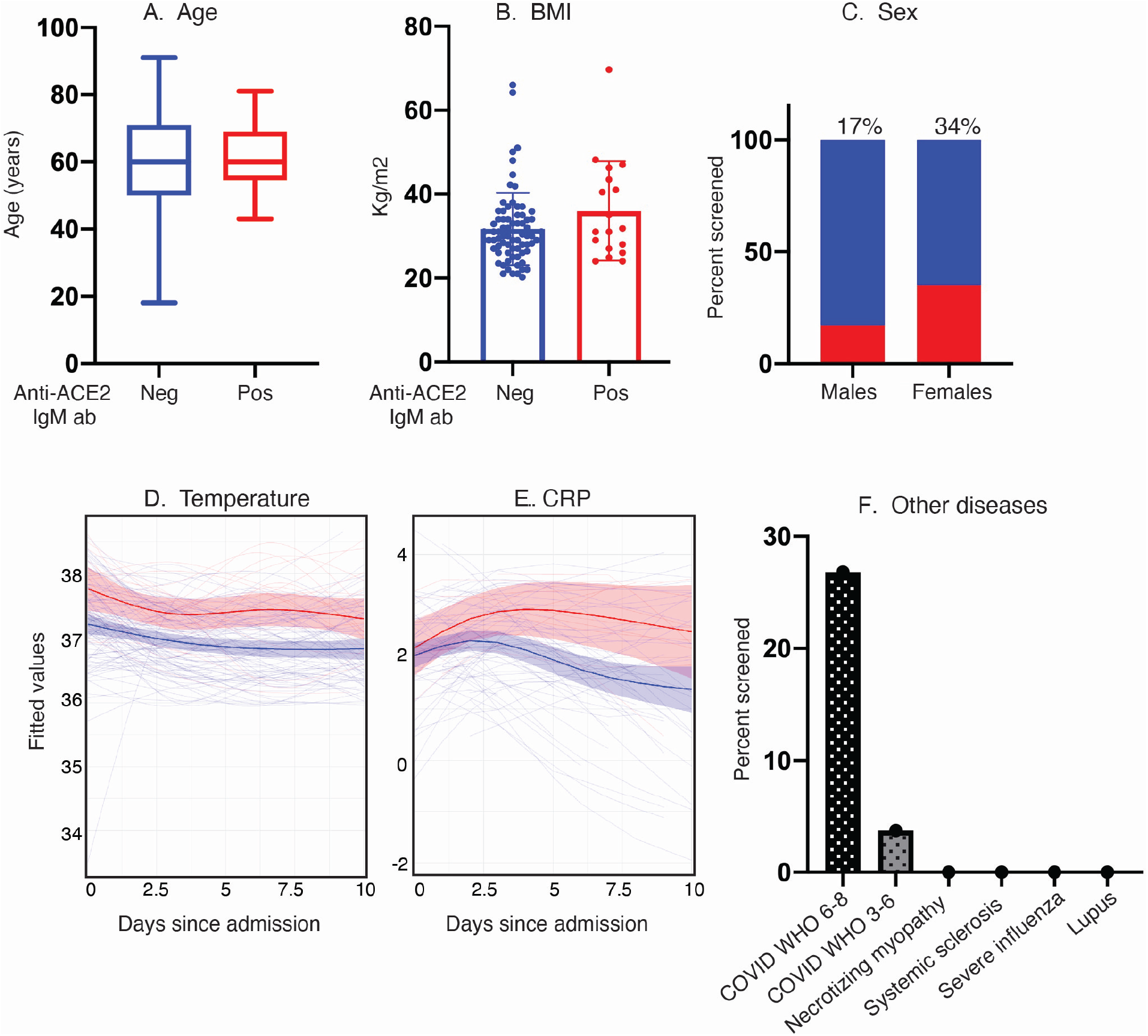
Clinical features of anti-ACE2 IgM-positive COVID-19 patients compared to those that do not have these antibodies. A-E: Age, BMI, sex, temperature and CRP levels were compared between the anti-ACE2 IgM-positive and negative COVID patient groups. Red and blue colors denote anti-ACE2 IgM-antibody positive and negative status, respectively. Box plots show median, 25^th^ and 75^th^ percentiles, and whiskers min to max. **Fig. 2D, E. IgM anti-ACE2 patients have higher average body temperature and CRP measurements beginning early after hospital admission**. The IgM anti-ACE2-positive group had statistically significantly higher average temperatures and CRP levels over the first 10 days of hospitalization than the IgM-negative group (p = 0.0001 and 0.02, respectively). Analyses in both 2D and 2E use linear mixed-effects model Wald test with 4 degrees of freedom (see statistical methods. **2F**: Anti-ACE2 IgM antibodies are detected in COVID-19 patients but not in other infectious and autoimmune disease controls.

### Longitudinal analysis of anti-ACE2 IgM and IgG suggests a T-independent autoantibody response

Since IgM is the earliest isotype elaborated in immune responses, we pursued a longitudinal analysis of anti-ACE2 IgM on all positive patients for whom serum was available. This demonstrated several patterns (Fig 1B, and Supp. Fig 3): (i) In 3 patients (CV-117, CV-123, CV-128), sampling spanned the development of anti-ACE2 IgM (Fig.1B and Suppl Fig 3). In these cases, autoantibodies appeared at ∼10 days after admission, and around the time of clinical worsening and intubation. We have not captured sufficient numbers of events around this time to make a definitive statement about onset of antibodies, but they do not appear to significantly precede clinical worsening; (ii) Anti-ACE2 IgM were already elevated at the first time point assayed in most patients, where patients were already intubated; in 4 patients (CV-1, CV-58, CV-65, CV-126), levels remained stable over time (one example shown in Fig. 1B; additional examples in Supp Fig. 3); (iii) In a third group, anti-ACE2 IgM levels decreased over time (CV-113, CV-124 and CV-134 (Fig 1B); CV-3, CV-57, CV-64, CV-129, CV-140, CV-143 (Suppl Fig 3)).

**Figure 3:**
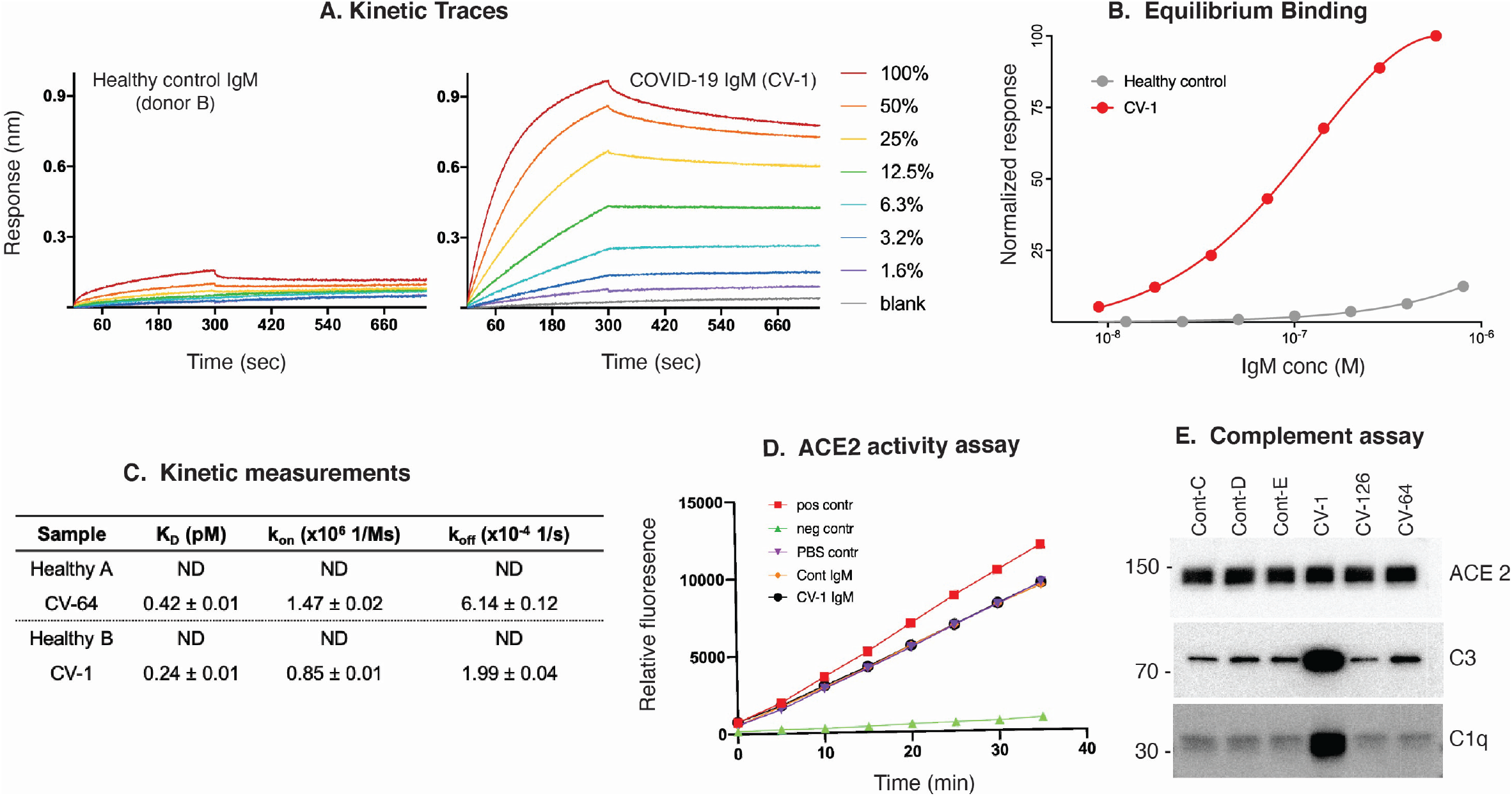
Properties of anti-ACE2 IgM antibodies. (A-C): Kinetic analysis. A: Kinetic traces of the binding interactions between immobilized human ACE2 and purified IgM, as determined by biolayer interferometry. Percentages represent twofold dilutions of IgM from patient CV-1 and Control B. B: Equilibrium binding titrations. Normalized responses at the indicated concentrations of purified IgM from the donors shown in (A) are plotted. C: Quantitation of the data obtained in A&B, and a separate patient and control shown in Supp. Fig 5A&B. **D: Anti-ACE2 IgM antibodies do not inhibit ACE2 activity**. ACE2 activity, in the presence or absence of IgM from patient CV-1 or Control B, was measured using a fluorescent substrate in a time course assay. The positive control was ACE2 alone, and the negative control was ACE2 plus ACE2 inhibitor (see Suppl Fig.5D for data obtained from another patient and control). **E: Complement activation induced by IgM antibodies to ACE2**. Dynabeads containing immune complexes of ACE2 and purified IgM from controls or anti-ACE2-positive COVID-19 (CV) patients were incubated with human complement. Deposition of C1q and C3 was visualized by immunoblotting. ACE2 is shown as a loading control. Markedly enhanced C1q binding in CV-1 observed in 3 separate experiments.

Overall, our studies captured multiple individuals where IgM levels decreased over time (Fig. 1B, Supp. Fig. 3). In T cell-dependent immune responses, this generally occurs at the time of class switching to IgG. We therefore examined whether decreasing IgM levels over time were associated with increasing anti-ACE2 IgG levels at later time points. In one patient (CV-1), both IgG and IgM were present at the earliest point and remained constant over time (Fig. 1B). In 8 anti-ACE2 IgM-positive patients, we observed a decrease of anti-ACE2 IgM to ∼50% over time; all these patients were IgG-negative and remained so.

Multiple groups have noted that high levels of anti-SARS-CoV-2 spike (S) protein IgG occurring at the time of hospital admission are associated with more severe disease in COVID-19 (e.g. ref (*18*)). We assayed antibodies to S-protein by ELISA, and anti-S and -RBD antibodies by the CoronaChek assay (Supp. Fig. 4). The mean ODs of anti-S antibodies were significantly higher in patients with severe compared to mild COVID-19 (0.68 +/- 0.48 vs 0.23 +/- 0.33; mean +/- SD, P<0.0001; Supp. Fig. 4A). Anti-S IgG levels were also significantly increased in anti-ACE2 IgM-positive COVID-19 patients compared to anti-ACE2 IgM-negatives (median 0.55 vs 0.14; p=0.028; Mann-Whitney test; Supp. Fig 4B). Using the CoronaChek assay, 8/8 (100%) of anti-ACE2 IgM-positive patients had a positive anti-ACE2 IgG result, compared to only 31/58 (53.4%) of anti-ACE2 IgM-negative patients (p=0.017, Fisher’s exact test; Supp. Fig 4C). Since anti-ACE2 IgM-positive patients have evidence of a robust anti-viral IgG response, failure of the anti-ACE2 IgM to isotype switch to IgG is not a general feature of anti-ACE2 patients. Instead, these data strongly suggest that the anti-ACE2 IgM immune response is not predominantly driven by T cells (either anti-viral or autoreactive), but rather represents a T-independent antibody response induced by SARS-CoV-2 infection. Such T-independent responses generally arise from B1 or marginal zone B cells(*19*),(*20*), and we suggest that such cells are the likely origin of this response. The finding of strikingly expanded circulating plasmablasts in severe COVID-19(*21*),(*4*),(*22*), a response which is oligoclonal with some clones that have not undergone somatic mutation, is consistent with this mechanism. An intriguing possibility is that a robust neutralizing anti-S IgG response induces an anti-idiotype IgM response, which also cross-reacts with ACE2, the spike protein receptor. The finding that anti-ACE2 IgM only appear after about 8-10 days after admission, where anti-S responses are well-established, is consistent with this possibility. The enhanced clinical inflammatory features (increased body temperature and CRP levels) that occur in anti-ACE2 IgM patients early after admission provides additional support for this hypothesis (Fig. 2D & E), as it suggests that the amplification to severe disease may begin significantly before decompensation(*2*).

**Figure 4:**
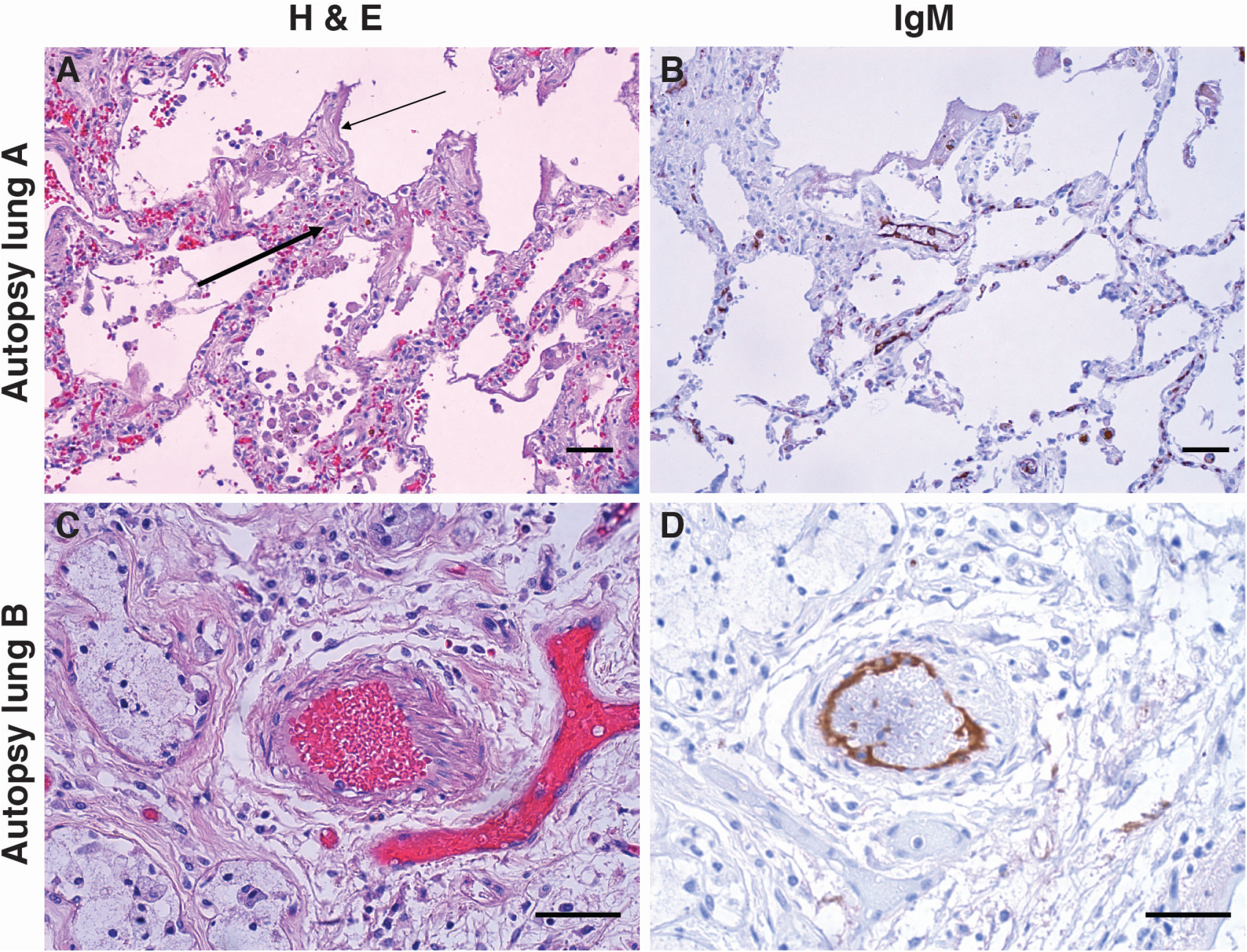
IgM deposition on endothelium in COVID-19 lung. Lung paraffin sections from two autopsy patients (lung A, upper panels; lung B, lower panels) were stained with hematoxylin and eosin (A & C) or with an anti-IgM antibody (B & D). A: A section of the left upper lobe of the lung shows a widened interstitium with capillaries showing reactive endothelium (thick arrow). There are hyaline membranes lining alveolar spaces (thin arrow), consistent with the exudative phase of diffuse alveolar damage (acute lung injury). B: Anti-IgM immunohistochemical staining of the same tissue highlights capillary endothelium in that area. C: A small artery of a bronchiole stained with hematoxylin and eosin, with (D) endothelial staining for anti-IgM. Size bars represent 50 microns.

The clinical efficacy of steroids in patients with severe COVID-19(*13*),(*23*) prompted us to examine whether any of the anti-ACE2 IgM-positive patients had been treated with steroids, and whether there was any relationship to subsequent IgM levels. None of the 5 anti-ACE2 IgM-positive individuals who died were treated with steroids for more than 2 days. Of the 13 patients that were ventilated but survived, 6 were treated with steroids for more than 2 days, and there was a temporal association of decreased anti-ACE2 IgM levels with steroid treatment in 3 patients where appropriate samples were available (Fig. 1B and Supp Fig. 3). The data suggest that anti-ACE2 IgM might be a dynamic biomarker, and worthy of study in a prospective cohort where the effects of steroids and other immune therapies can be rigorously addressed.

### Properties of IgM autoantibodies recognizing ACE2

We next pursued additional analysis of the anti-ACE2 IgM binding properties using a different source of ACE2 antigen, and a different assay format. IgM purified from 2 patients with high titer ACE2 antibodies and 2 healthy controls were analyzed via biolayer interferometry. Patient IgM binding to immobilized ACE2 was saturable, with apparent K_D_ values (Fig 3C) of 0.11μM (CV-1) and 3.6 μM (CV-64), whereas IgM from healthy controls did not exhibit measurable binding to ACE2 (Fig. 3A-C and Supp. Fig.5, panels A-C). The reported K_D_ values provide a ceiling for these measurements; since the purified IgM is a mixture of antibodies against various targets, the actual ACE2 affinities for individual IgM clones are presumably higher. These data are consistent with antibodies that have not undergone affinity maturation, which are known to have low affinity (high nanomolar to micromolar range) but benefit from avidity effects (*24*).

Since hypomorphic ACE2 function has been associated with severity in ARDS, we investigated whether purified IgM from COVID-19 patients affected the catalytic function of ACE2 against a fluorogenic substrate. The purified IgM used above in binding assays had no effect on ACE2 activity (Fig. 3D and Supp. Fig5C).

IgM antibodies are mainly found in the circulation, where they are the most effective isotype at activating the classical complement cascade at surfaces expressing their cognate antigens(*25*). We found that IgM antibodies with high affinity binding to ACE2 (i.e. CV-1) consistently activated complement upon antigen binding (Fig. 3E). In some experiments, IgM purified from CV-64 and CV-164 behaved similarly to CV-1, although the magnitude of the effect was decreased (Supp. Fig 5D). These data suggest that IgM antibodies recognizing ACE2 play a role in the widespread complement pathway activation observed in COVID-19 patients (*7*)(*26*).

Recent autopsy studies in COVID-19 have demonstrated a striking series of findings in the lung of COVID-19 patients(*9*),(*27*). In addition to diffuse alveolar damage and perivascular infiltrating lymphocytes, there were striking angiocentric features in COVID-19 lungs, including severe endothelial injury associated with membrane disruption and ACE2 expression, widespread microangiopathy with occlusion of capillaries, and new vessel growth. In order to define whether there was any in vivo evidence of IgM deposition in the lungs of patients with COVID-19, specimens from 23 autopsies were stained with anti-human IgM. Serum from the biorepository was only available in 7 of these patients; they were all negative for serum anti-ACE2 IgM, and had no IgM staining in the lung. Among the remaining patients, 4 (25%) had evidence of staining with anti-IgM, with staining observed in blood vessels and capillaries in 2 patients (Fig. 4). Another patient had staining of a lymphatic channel, and the final patient had staining that was alveolar in pattern. Vascular endothelium appeared reactive, mostly without accompanying inflammatory infiltrates. The finding of examples of endothelial IgM deposition in COVID-19 lung demonstrates that an endothelial cell target with similar distribution to ACE2 (*9*),(*27*) is recognized by IgM in a subgroup of COVID-19 patients with fatal disease. Defining whether these IgM molecules recognize ACE2 or another endothelial antigen is a high priority.

These studies demonstrate that anti-ACE2 IgM arise in the context of severe COVID-19, likely predominantly as a T-independent antibody response. This immune response is of potential pathogenic significance through binding to the surface of endothelial cells, activating the classical complement cascade, and initiating an inflammatory response. These mechanisms are potentially amenable to several readily available treatments, particularly short duration anti-inflammatory therapy (e.g. steroids and IVIG therapy (*28*),(*29*),(*30*) and potentially inhibitors of complement or therapies targeting T-independent antibody generation). It is noteworthy that mortality of the dermatopulmonary phenocopy associated with MDA5 autoantibodies appeared to be substantially decreased by steroids, IVIG and calcineurin inhibitor treatment (*31*)(*32*), although controlled trials have not been possible in this rare phenotype. Since ACE2 autoantibodies have features of T-independent responses, this may provide an important opportunity to use focused, short-term immune-focused therapies in severe COVID-19 (consistent with the dexamethasone results(*13*)) rather than the deeper immunosuppression needed for T cell-driven processes.

In contrast to the recently described genetic and preexisting autoimmune factors that predispose to severe COVID-19 (i.e., inborn errors of type I IFN immunity and anti-IFN autoantibodies (*33*)), this study adds a unique biomarker that results from SARS-CoV-2 infection and is strongly associated with severe clinical outcomes in patients with COVID-19. The 27% of those with critical disease who develop IgM to ACE2 (more women, antibodies follow infection) are almost certainly non-overlapping with the 10% of severe COVID-19 patients who have prexisiting IgG to Type I IFNs (autoantibodies are IgG, almost exclusively in males, and precede infection) (33). Together with an additional 2-3% have inborn errors of type I IFNs (33), these constitute about 40% of severe COVID-19 patients. Another major endophenotype in severe COVID-19 appears to encompass patients with immunosenescence, with blunted CD8 responses, which is enriched in the elderly (*34*)(*35*). These endophenotypes, driven by distinct mechanisms, having actionable markers and accounting for a substantial fraction of severe COVID-19, will likely benefit from both shared and distinct therapeutic approaches. Rapidly defining additional mechanistically-anchored groups in severe COVID is a high priority.

## Data Availability

Patient data and autoantibody data are stored in JH-CROWN, hosted on the Johns Hopkins Precision Medicine Analytics Platform. Data or materials derived from human samples may be requested subject to any underlying restrictions on such data or samples, and will require material transfer and data use agreements through Johns Hopkins University.

## Acknowledgements

The data and specimens utilized were part of JH-CROWN: The COVID-19 PMAP Registry, and the Johns Hopkins COVID-19 Remnant Specimen Repository, respectively. These resources are based on the contribution of many patients and clinicians, and were funded by the University and Hopkins inHealth, the Johns Hopkins Precision Medicine Program. Work with influenza samples was supported in part by the US Department of Health and Human Services Biomedical Advanced Research and Development Authority (BARDA; agreement number IDSEP160031-01-00) and the National Institute of Allergy and Infectious Diseases Contract HHSN272201400007C awarded to the Johns Hopkins Center for Influenza Research and Surveillance (JHCEIRS) at the Johns Hopkins University. We thank Dr. Christopher Karp from the Bill & Melinda Gates Foundation, Dr. Stephen Desiderio and Dr. Paul Rothman from Johns Hopkins for numerous helpful discussions. The content of this paper is solely the responsibility of the authors, and does not represent the official views of the NIH.

## Funding

Funding for these studies were provided by the Bill & Melinda Gates Foundation (BMGF), Gates Philanthropy Partners, the Donald and Dorothy Stabler Foundation, and the Jerome L. Greene Foundation. This study was supported in part by NIH grants R01 AR073208 (LCR), R01 AR069569 (FA), NIH IRACDA program (5K12GM123914-03, EKL) and the Division of Intramural Research, National Institutes of Allergy and Infectious Diseases (OL). The Rheumatic Diseases Research Core Center, where the ELISA assays were performed, is supported by NIH P30-AR070254.

## Author Contributions

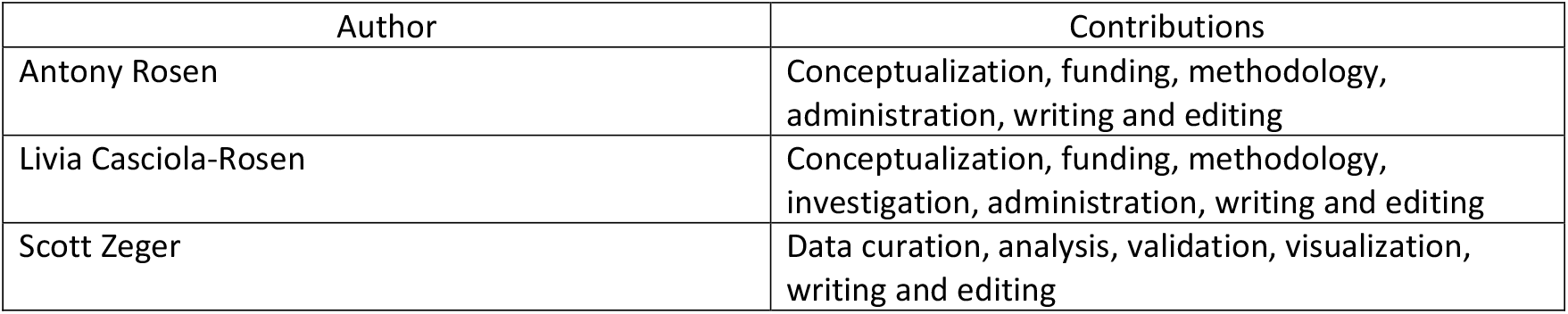
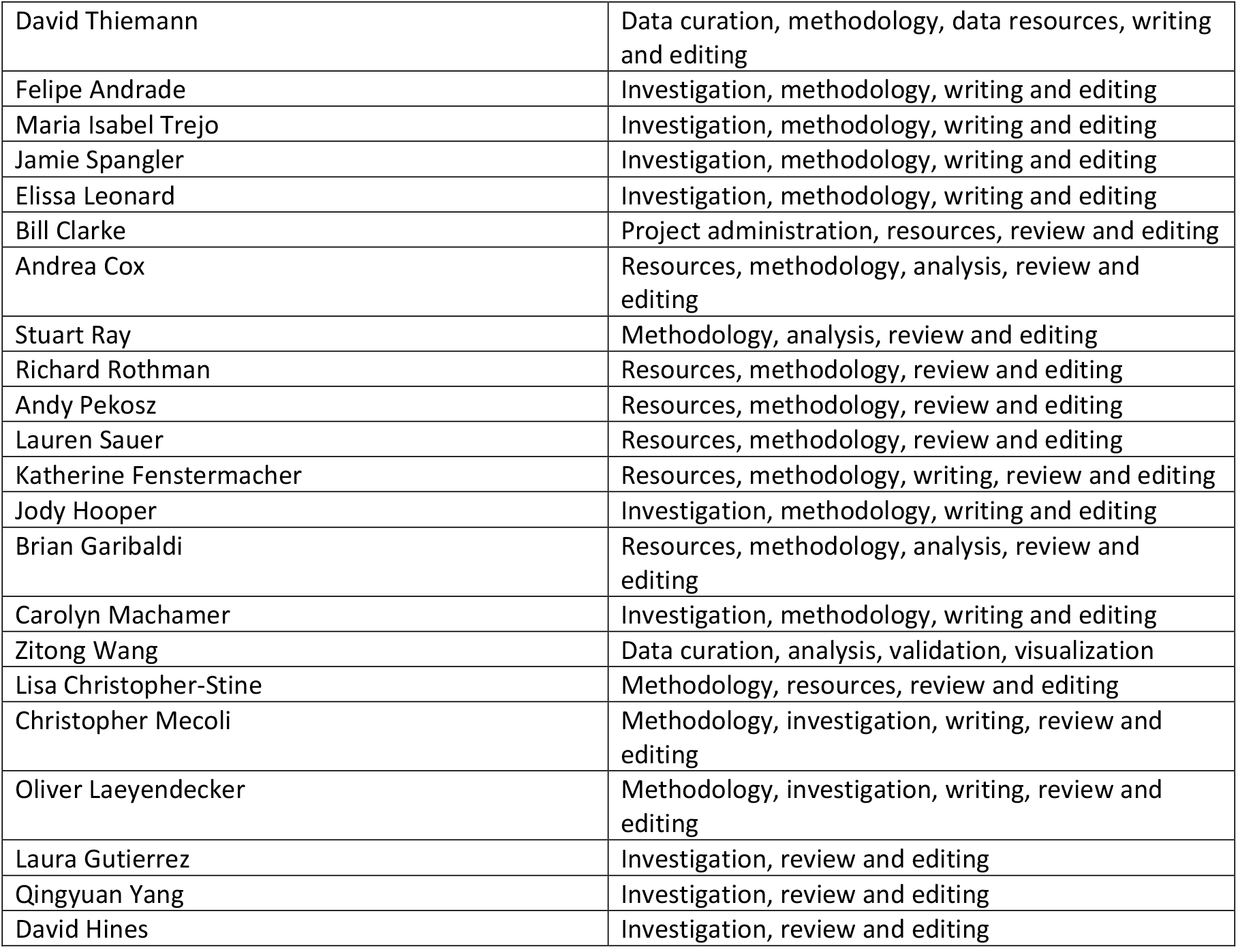

| Author | Contributions |
| --- | --- |
| Antony Rosen | Conceptualization, funding, methodology, administration, writing and editing |
| Livia Casciola-Rosen | Conceptualization, funding, methodology, investigation, administration, writing and editing |
| Scott Zeger | Data curation, analysis, validation, visualization, writing and editing |
| David Thiemann | Data curation, methodology, data resources, writing and editing |
| Felipe Andrade | Investigation, methodology, writing and editing |
| Maria Isabel Trejo | Investigation, methodology, writing and editing |
| Jamie Spangler | Investigation, methodology, writing and editing |
| Elissa Leonard | Investigation, methodology, writing and editing |
| Bill Clarke | Project administration, resources, review and editing |
| Andrea Cox | Resources, methodology, analysis, review and editing |
| Stuart Ray | Methodology, analysis, review and editing |
| Richard Rothman | Resources, methodology, review and editing |
| Andy Pekosz | Resources, methodology, review and editing |
| Lauren Sauer | Resources, methodology, review and editing |
| Katherine Fenstermacher | Resources, methodology, writing, review and editing |
| Jody Hooper | Investigation, methodology, writing and editing |
| Brian Garibaldi | Resources, methodology, analysis, review and editing |
| Carolyn Machamer | Investigation, methodology, writing and editing |
| Zitong Wang | Data curation, analysis, validation, visualization |
| Lisa Christopher-Stine | Methodology, resources, review and editing |
| Christopher Mecoli | Methodology, investigation, writing, review and editing |
| Oliver Laeyendecker | Methodology, investigation, writing, review and editing |
| Laura Gutierrez | Investigation, review and editing |
| Qingyuan Yang | Investigation, review and editing |
| David Hines | Investigation, review and editing |

## Competing interests

Antony and Livia Casciola-Rosen are listed as inventors on a patent application filed by Johns Hopkins University that encompasses aspects of this publication. All other authors declare no competing interests.

## Data and Materials availability

Patient data and autoantibody data will be stored in the JH-CROWN registry, which is housed on the Johns Hopkins Precision Medicine Analytics Platform. Requests for data or materials derived from human samples may be made available, subject to any underlying restrictions on such data and samples, and will require material transfer and data use agreements through Johns Hopkins University.

## Materials and Methods

### Patient data and serum samples

The study cohort was defined as inpatients who had: 1) a confirmed diagnosis of COVID-19; 2) survival to death or discharge; and 3) remnant specimens in the Johns Hopkins COVID-19 Remnant Specimen Biorepository, an opportunity sample that includes 59% of Johns Hopkins Hospital COVID-19 patients and 66% of patients with length of stay >=3 days. Diagnosis of COVID-19 was defined as detection of SARS-CoV-2 using any PCR test with an Emergency Use Authorization from the US Food and Drug Administration. Selection and frequency of other laboratory testing were determined by treating physicians. The primary clinical data source was JH-CROWN, a Johns Hopkins Medicine COVID-19 registry that integrates all clinical data for COVID-19 patients, including demographics, medical history, comorbid conditions, symptoms, medications, laboratory results, medical images, and comprehensive bedside flowsheet data, including vital signs, respiratory events, and intravenous medication titration (*2*).

Patient outcomes were defined by the World Health Organization (WHO) COVID-19 disease severity scale. The WHO scale is an 8-point ordinal scale ranging from ambulatory (1=asymptomatic, 2=mild limitation in activity), to hospitalized with mild-moderate disease (3=room air, 4=nasal cannula or facemask oxygen), hospitalized with severe disease (5=high flow nasal canula (HFNC) or non-invasive positive pressure ventilation (NIPPV), 6=intubation and mechanical ventilation, 7=intubation and mechanical ventilation and other signs of organ failure (hemodialysis, vasopressors, extracorporeal membrane oxygenation (ECMO)), and 8=death. For this study we combined adjacent WHO classes, dividing the inpatient population into two groups according to maximum WHO severity: patients who did not require mechanical ventilation (WHO class 3-5); those who required mechanical ventilation with or without additional support, such as intravenous pressors, continuous renal replacement therapy (CRRT) and/or extracorporeal membrane oxygenation (ECMO) who survived (WHO classes 6-7) or died (WHO class 8). Serum samples were selected for timing within 24 hours of onset of the maximum WHO class; when multiple samples were available, the specimen closest to the WHO class onset was used. The initial analysis used a random sample of 12-20 unique patient specimens from each of the 4 classes meeting the criteria above (depending upon specimen availability for the clinical class). To determine biomarker trajectory, we analyzed an expanded cohort of patients who had 3-4 consecutive sera per patient across the course of their hospitalization. Patient selection was determined solely by specimen availability. Where available, additional serum for anti-ACE2 IgM-positive individuals was requested from the remnant biorepository. These studies were approved by the JHU Institutional Review Board (IRB 00251725, IRB 00256018, 00256547), with a waiver of consent because all specimens and clinical data were de-identified by the Core for Clinical Research Data Acquisition of the Johns Hopkins Institute for Clinical and Translational Research; the study team had no access to identifiable patient data. Patient numbers per analysis are denoted in the figure legends.

#### Disease and healthy control sera

Three autoimmune disease control cohorts consisted of the following. (i) Sera from N=25 patients with SLE from the Johns Hopkins Lupus Cohort. (ii) Sera from N=13 patients diagnosed with systemic sclerosis after evaluation at the Johns Hopkins Scleroderma Center. (iii) N=15 patients with necrotizing myopathy defined by a positive anti-HMGCR antibody status evaluated at the Johns Hopkins Myositis Center. Serum from N=30 patients with influenza diagnosed using the Cepheid Xpert Xpress Flu/RSV assay in the Johns Hopkins Emergency departments or in-patient units, were studied. 11 were evaluated in the ED and discharged to outpatient care; an additional 19 patients were hospitalized, and required oxygen therapy or assisted ventilation (5). Sera from N=30 adult healthy control individuals were also studied. Informed consent for these samples was obtained following protocols approved by the JHU Institutional Review Board (NA_00039294, NA_00039566, #NA00007454, IRB00066509, IRB00091667).

At the initiation of this study, we were struck by the similar clinical presentation of severe COVID-19 to a dermatopulmonary syndrome, characterized by skin rash, rapidly progressive interstitial lung process with frequent progression to a need for ventilatory support, and a unique vasculopathic phenotype including cutaneous ulcers and digital ischemia (*17*)(*36*)(*37*). This syndrome has been associated with IgG autoantibodies to melanoma differentiation-associated 5 (MDA5)(*38*). In its fulminant form, this can be viewed as a phenocopy of severe COVID-19, with a high mortality in the absence of treatment with steroids, IVIG, or calcineurin inhibition (*31*)(*32*). Serum was available to us from a 42 year-old patient with this MDA5-associated syndrome, who developed symptoms of weakness, rash, fevers, and dyspnea in October of 2011. Her clinical course stabilized with immunosuppression consisting of corticosteroids, tacrolimus, and rituximab over the ensuing 8 years. Informed consent was obtained at presentation following protocols approved by the Johns Hopkins Institutional Review Board #NA00007454. Strikingly, this index patient had IgM and IgG autoantibodies against ACE2; her serum served as the reference calibrator on all ELISA plates. A study to understand the prevalence and relevance of these ACE2 autoantibodies in anti-MDA5-positive dermatomyositis-like disease and other rheumatic syndromes characterized by severe lung disease is currently ongoing.

#### Anti-ACE2 and -SARSCoV2 spike ELISA assays

ELISA plate wells were coated overnight with 50 ng of purified protein (recombinant human ACE2 from Abcam, cat # ab151852; SARSCoV2 spike protein S1 subunit from Sino Bio cat # 40591-V08B1) diluted in PBS. For each serum assayed, 2 wells were coated with protein (duplicate readout), and an adjacent well was incubated overnight with PBS only (to determine background specific to each sample tested). *Anti-ACE2 IgM ELISA*: Wells were washed with PBS plus 0.1% Tween (PBST), and subsequently blocked with 3% milk/PBST. Primary antibody incubations were routinely performed by diluting sera 1:200 in 1% milk/PBST overnight at 4°c. For the area under the curve plots (shown in Suppl. Fig 1B), serial serum dilutions ranging from 1:100 to 1:3,200 were used for the ELISA assays. Wells were then washed with PBST, followed by incubation with HRP-labeled anti-human IgM (Heavy chain-specific; Jackson ImmunoResearch cat # 109-035-043) diluted 1:5000 in 1% milk/PBST (1 hour, room temperature). Color was developed with SureBlue peroxidase reagent (KPL). Reactions were terminated by adding HCl, and absorbances were read at 450 nM. The same anti-ACE2 IgM-positive reference serum was included on each plate assayed and all absorbances were calibrated relative to this reference serum. *Anti-ACE2 IgG ELISA*: was performed as described for anti-ACE2 IgM antibodies, with the following modifications. The concentration of Tween in PBST was 0.5%. Blocking was performed with 5%BSA/PBST, and sera and secondary antibodies were diluted with 1% BSA/PBST. The secondary antibody was HRP-labeled anti-human IgG (Jackson ImmunoResearch cat # 109-036-088), diluted 1:10,000. The cutoff for assigning anti-ACE2 IgM and IgG antibody positivity was determined by assaying sera from 30 healthy controls. The mean +3 SD of these values (0.340 and 0.187 calibrated OD units for anti-ACE2 IgM and IgG antibodies, respectively) was taken as the cutoff for each. The anti-ACE2 ELISA was validated by (i) blotting purified recombinant human ACE2 and (ii) using a second source of recombinant human ACE2 purchased from another vendor (Sino Biological, cat # 10108-H08H). *Anti-SARSCoV2 spike IgG ELISA* – These assays were performed as described for anti-ACE2 IgG ELISAs, with the following modifications. Sera were assayed at a 1:1,200 dilution, and the primary antibody incubation was performed for 1 hr at room temperature.

#### CoronaChek assay

The CoronaChek serologic lateral flow assay (Hangzhou Biotest Biotech Co, Ltd., Hangzhou China) detects M (IgM) and G (IgG) antibodies to the spike protein and receptor binding domain of SARS-CoV-2. Studies on positive and negative control specimens from Maryland demonstrated: sensitivity of 95%, (95%CI 83%, 99%) in convalescent plasma donors an average of 50 days post symptom onset; sensitivity of 100% (95%CI 89%, 100%) in PCR confirmed hospitalized individuals 15 days after symptom onset; specificity of 100% 95% CI 94%, 100%) in pre-pandemic patients infected with rhinoviruses and other coronaviruses.

#### Purification of IgM from patient serum

Following the manufacturer’s instruction, 0.5 mL of POROS CaptureSelect™ IgM Affinity Matrix (Thermo Fisher Scientific) was equilibrated with 10 column volumes (CV) of phosphate-buffered saline (PBS) pH7.2 in a Poly-Prep® chromatography column (Bio-Rad). Patient serum samples (400 µL) were diluted 1:10 in PBS pH 7.2, filtered via centrifugation at 12,000×g using 0.45 µm spin filters (EMD Millipore), and loaded onto the column. The column was washed twice with 5 CV of PBS pH 7.2. Bound IgM protein was eluted with 5 CV of 0.1 M glycine, pH 3. The eluted IgM was immediately neutralized with 0.1 CV of 1 M Tris-HCl (pH 8).

The eluted IgM was exchanged into PBS and concentrated to match the original serum volume using Amicon 30 kDa molecular weight centrifugal filters (EMD Millipore). The 280 nm absorbance of the purified IgM was measured to calculate the IgM concentration, using the extinction coefficient for pentameric human IgM.

#### Biolayer interferometry analysis of ACE2/IgM interaction

Biolayer interferometry was performed using an Octet RED96 instrument (Molecular Devices) to measure the interaction of purified IgM from patient serum to ACE2. Wells of a black flat-bottom polypropylene plate (Corning) were loaded with the following samples: 50 nM biotinylated human ACE2 (Sino Biological, 10108-H08H-B); twofold dilutions of purified patient IgM; PBSA (PBS pH 7.2 containing 0.1% bovine serum albumin [BSA]); and regeneration buffer (0.1 M glycine, pH 3). All samples were centrifuged at 12,000×g through a 0.45 μm filter device (EMD Millipore), and buffers were vacuum filtered using a 0.22 µm membrane (EMD Millipore). ACE2 and the IgM samples were diluted in PBSA. ACE2 was loaded onto hydrated streptavidin (SA) biosensor tips (Molecular Devices), and baseline measurements were collected in PBSA. Binding kinetics were then measured by submerging the ACE2-coated biosensors in wells containing twofold serial dilutions of each patient IgM sample for 300 s (association) followed by submerging the biosensor in wells containing only PBSA for 450 s (dissociation). Tips were regenerated via exposure to regeneration buffer. Analysis and kinetic curve fitting (assuming a 1:1 binding model) was conducted using Octet Data Analysis HT software version 7.1 (Molecular Devices). Normalized equilibrium binding curves were obtained by plotting the response value after the 300 s association phase for each sample dilution and normalizing to the maximum value. Equilibrium curves were fitted to a single logistic model using a non-linear regression algorithm in GraphPad Prism software.

#### ACE2 activity assay

ACE2 activity was measured using a kit from BioVIsion (K897). Purified IgM (5 μg) or ACE2 inhibitor was preincubated with ACE2 in white Costar 96-well plates for 20 min at RT, followed by addition of fluorogenic ACE2 substrate as per the manufacturer’s protocol. PBS made up 20% of the assay volume for CV-1 IgM and 10% for CV-64 IgM (due to lower protein concentration of the CV-1 IgM). Thus, a PBS control was included for each assay. The positive control contained only ACE2 and substrate, and the negative control was ACE2 plus ACE inhibitor and substrate. Fluorescence was measured every 5 min after substrate addition in a BMG Labtech FLUOstar Omega plate reader, with excitation at 355 nm and emission at 460 nm. Fluorescence values for wells containing no ACE2 (blank) were subtracted from the values shown.

#### Complement activation assay

20 μl Dynabeads M-270 streptavidin (Thermo) were coated with 250 ng of biotinylated ACE2 purchased from either ACROBiosystems (Cat# AC2-H82E6) and SinoBiological (Cat: #10108-H08H-B). These were then incubated with 0.5μg purified IgM (see above) diluted in 200 μl NP40 Buffer A (20 mM Tris pH 7.4, 150 mM NaCl, 1 mM EDTA) containing 1% BSA. After 2 hrs at RT, the beads were washed twice in Buffer A and once in gelatin veronal buffer (GVB, Comptech). Human serum was added as the source of complement (1:50 dilution) to reach a final volume of 100 μl in GVB. After 1 hr at 37°c, the beads were washed in Buffer A, and boiled in gel application buffer. Samples were analyzed by electrophoresis on 12% SDS-polyacrylamide gels. ACE2, C3 and C1q were detected by immunoblotting (anti-ACE2, R&D systems Cat# AF933; anti-C3, Santa Cruz Cat# sc28294; anti-C1q, Comptech Cat# A200). Proteins were detected using horseradish peroxidase–labeled secondary antibodies (Jackson ImmunoResearch) and chemiluminescence. Images were acquired using a Protein Simple Fluorochem-M digital imager.

#### Immunohistochemistry

Autopsies of 23 patients infected with Sars-CoV-2, documented by PCR on a pre or postmortem nasopharyngeal swab, were examined. Autopsies were consented for and performed on the clinical service with complete examination of chest organs and in-situ sampling of remaining organs and tissues, with histology performed on all sites. Lung paraffin sections from COVID-19 autopsy patients were either stained with Hematoxylin and Eosin, or processed as follows. After deparaffinization and rehydration, the sections were immersed in antigen retrieval solution (DAKO) for 30 min at 98°c. For IgM staining, the sections were blocked with goat serum (30 minutes at room temperature), followed by incubation with horseradish peroxidase labeled goat anti-human IgM (Jackson ImmunoResearch, cat # 109-035-043) diluted 1:500. Visualization was performed with a liquid DAB substrate-chromagen system (DAKO) and the sections were counterstained with hematoxylin before mounting.

#### Statistical Methods

The clinical measures used in this analysis are from the JHM COVID-19 Crown Registry that is actively curated by a team of clinicians, informaticists, and statisticians to assure data quality. For repeated measures outcomes (e.g. temperature, CRP, BMI) data was checked by making spaghetti plots (*39*) and visually checking the consistency of observations over time within an individual. The other main source are laboratory measurements of immune status (e.g. IgM or IgG antibodies) that are either binary indicators of presence/absence or absorbance levels as described in the immunoassay section.

To compare the rates of IgM antibody positivity between two subgroups, we estimated the ratio of the odds of positivity for one subgroup versus the other (odds ratio) and 95% confidence interval. Given the small numbers of patients in some comparator groups, we used a Fisher’s exact test of the null hypothesis that the rates were equal (odds ratio = 1). To compare means of continuous variables with roughly Gaussian distributions (determined using a quantile-quantile plot), we estimated the mean difference and its standard error and used an unpaired t-test of the null hypothesis that the two population means are equal. When we detected a large deviation from Gaussianity (for S protein IgG), a non-parametric test (Mann-Whitney) was used instead.

To compare the trajectory of clinical outcomes over time between IgM positive and IgM negative groups, we used a linear mixed effects model (*40*). Variables were transformed to the log-scale if their marginal distribution was more nearly symmetric after transformation. The fixed effects included an indicator variable for IgM-positive status a smooth function of time (natural cubic spline with 3 degrees of freedom) and their interaction. We assumed each person had a random intercept and random linear trend to account for the likely correlation among repeated observations on individuals. Given this specification, we estimated the smooth curve for the IgM positive and negative groups as well as their difference with 95% confidence intervals. We tested the null hypothesis that the two population time curves are the same (coefficients for main effect of IgM and interaction of IgM with time all equal 0) using a Wald test statistic that was compared to a Chi-square distribution with 4 degrees of freedom. The analysis was repeated using natural splines with 2 to 4 degrees of freedom to assure that the findings were not sensitive to these assumptions.

## SUPPLEMENTAL FIGURE LEGENDS

**Supplemental Table 1.** Demographics of the study population.

|  |  | N |
| --- | --- | --- |
| Total |  | 118 |
| **Demographics** |  |  |
| Age (years) | 60 (50-71) |  |
| Male gender | 56% | 66 |
| White race/ethnicity | 26% | 31 |
| Black race/ethnicity | 41% | 48 |
| Hispanic race/ethnicity | 23% | 27 |
| Asian race/ethnicity | 3% | 3 |
| Other | 8% | 9 |
| BMI (kg/m<sup>2</sup>) | 30.2 (26.0-34.9) |  |
| **Comorbidities** |  |  |
| Diabetes mellitus | 47% | 56 |
| Hypertension | 64% | 76 |
| Coronary artery disease | 24% | 28 |
| Congestive heart failure | 23% | 27 |
| Chronic lung disease | 26% | 31 |
| **<u>Maximum WHO class</u>** |  |  |
| Minimal oxygen | 28% | 34 |
| HFNC/NIPPV | 15% | 18 |
| Mechanical ventilation | 32% | 38 |
| Dead | 24% | 28 |

Continuous variables are median +/- interquartile range
Categorical variables are percentages

**Supplemental Fig 1:**
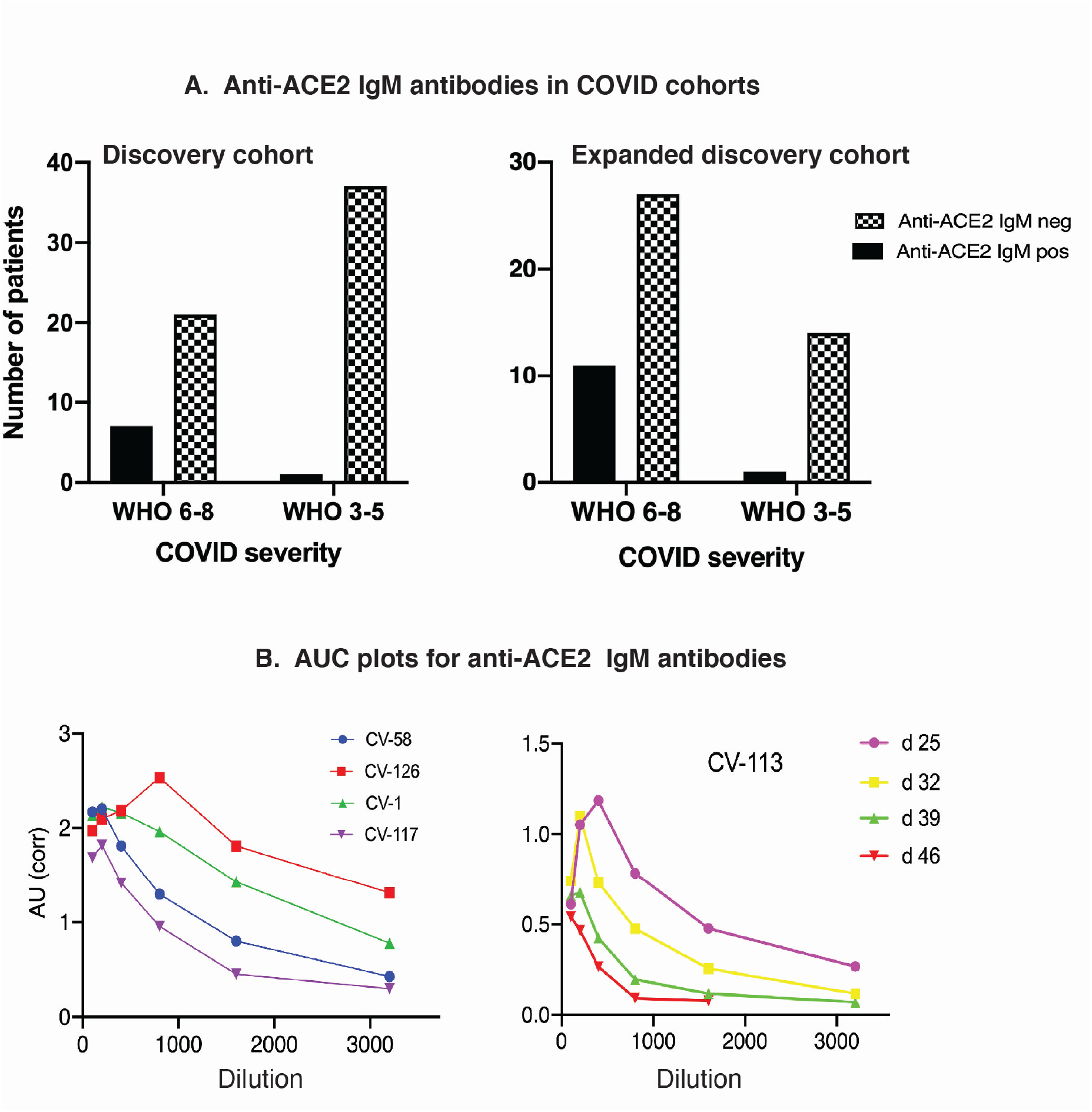
Anti-ACE2 IgM antibodies in COVID-19 patients. A. Anti-ACE2 IgM ELISAs were performed as described in the Methods section. In the Discovery cohort (left panel), 8/66 patients with COVID-19 were positive for anti-ACE2 IgM antibodies. Of these, 25% of the WHO 6-8 group were positive compared to 2.6% of the WHO 3-5 group (p=0.0084, Fisher’s exact test). An additional 52 COVID-19 patients were assayed (“Expanded discovery”, right panel); the frequency of anti-ACE2 IgM in these patients was similar to the initial group. Data from the combined cohorts (N = 118) is shown in Fig 1A. B. Anti-ACE2 IgM ELISAs were performed using serial serum dilutions (1:100 to 1:3,200 range). Data obtained from four different patients is shown in the left panel, each assayed using serum from a single bleed. Data from a fifth patient is shown in the right panel, using serum made from blood draws on 4 different days. Area under the curve (“AUC”) plots are shown in both panels.

**Supplemental Fig. 2:**
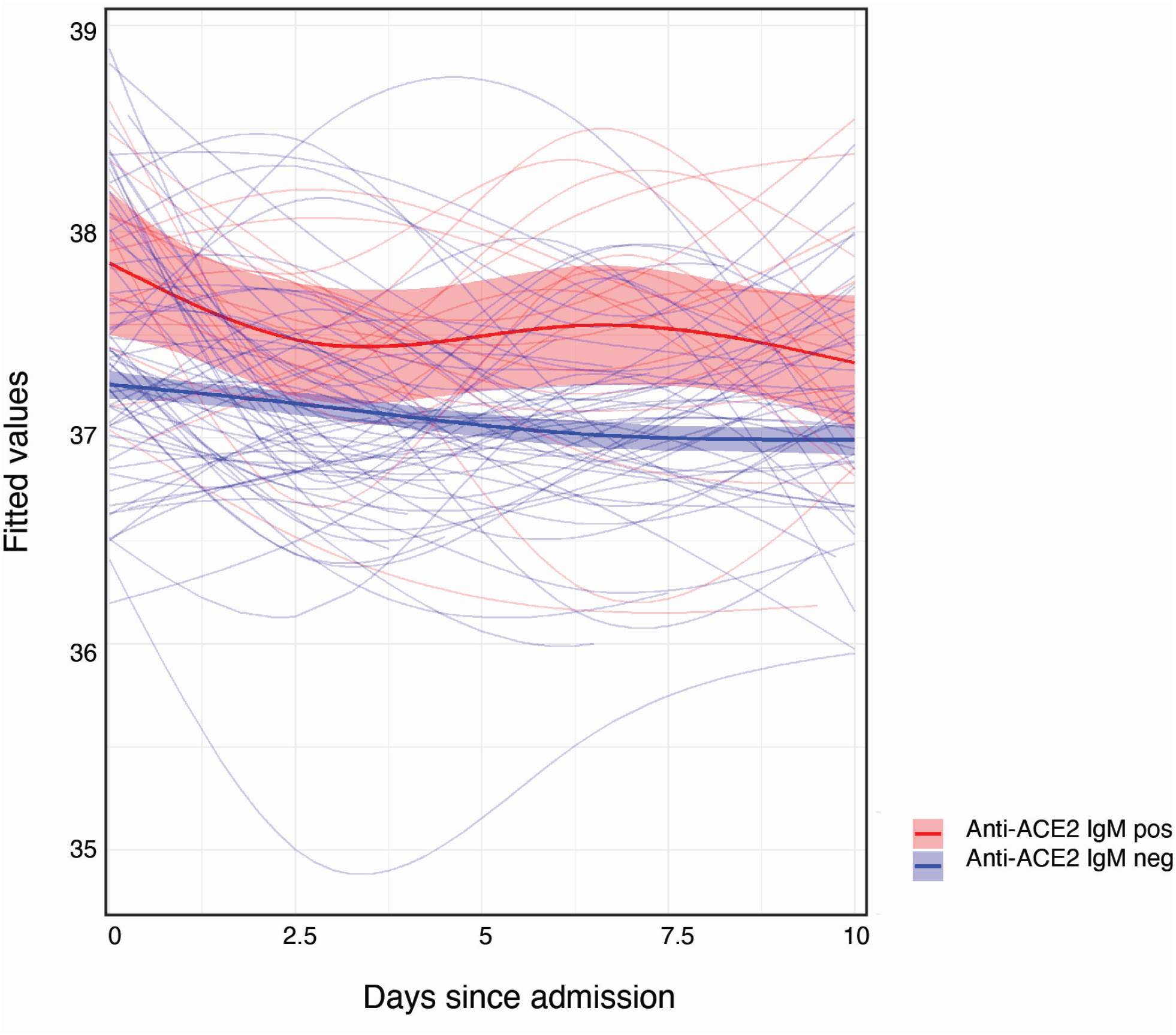
The higher average body temperature measurements in IgM anti-ACE2 patients are not a function of disease severity. IgM anti-ACE2-positive group had statistically significantly higher average temperatures over the first 10 days of hospitalization than the IgM-negative group (Fig. 2D). The analysis here is restricted to the severe IgM-positive patients compared to all severe COVID-19 patients from the CROWN Registry for whom IgM status was unknown. The results are unchanged, implicating the increased temperature as a function of IgM status rather than disease severity (IgM-positive: mean = 37.53, S^2^ = 0.64, N = 721 on M = 18 unique patients, IgM-unknown: mean = 37.11, S^2^ =0.59, N =14827 on M = 473 unique patients; chisq = 19.98, p = 0.0005 from linear mixed-effects model Wald test with 4 degrees of freedom.

**Supplemental Fig 3:**
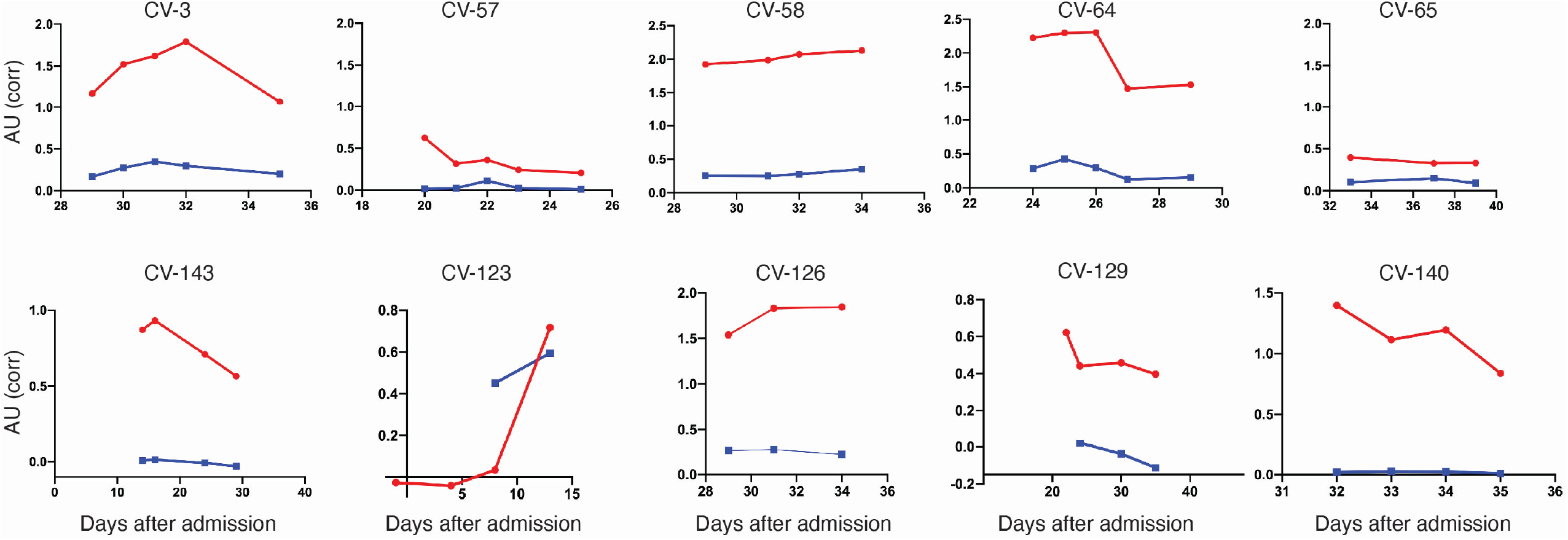
Longitudinal analysis of anti-ACE2 IgM antibodies in patients hospitalized with severe COVID-19. For all those anti-ACE2 IgM-positive patients with multiple banked sera, anti-ACE2 IgM and IgG antibodies were quantitated over time. Red and blue lines on each plot denote anti-ACE2 IgM and IgG antibodies, respectively. The following patients were on steroid treatment: CV-58 (days 20-24 and 29-36); CV-65 (days 26-28) and CV-129 (day 20 to beyond day 60). Additional examples are shown in Fig. 1B.

**Supplemental Fig. 4:**
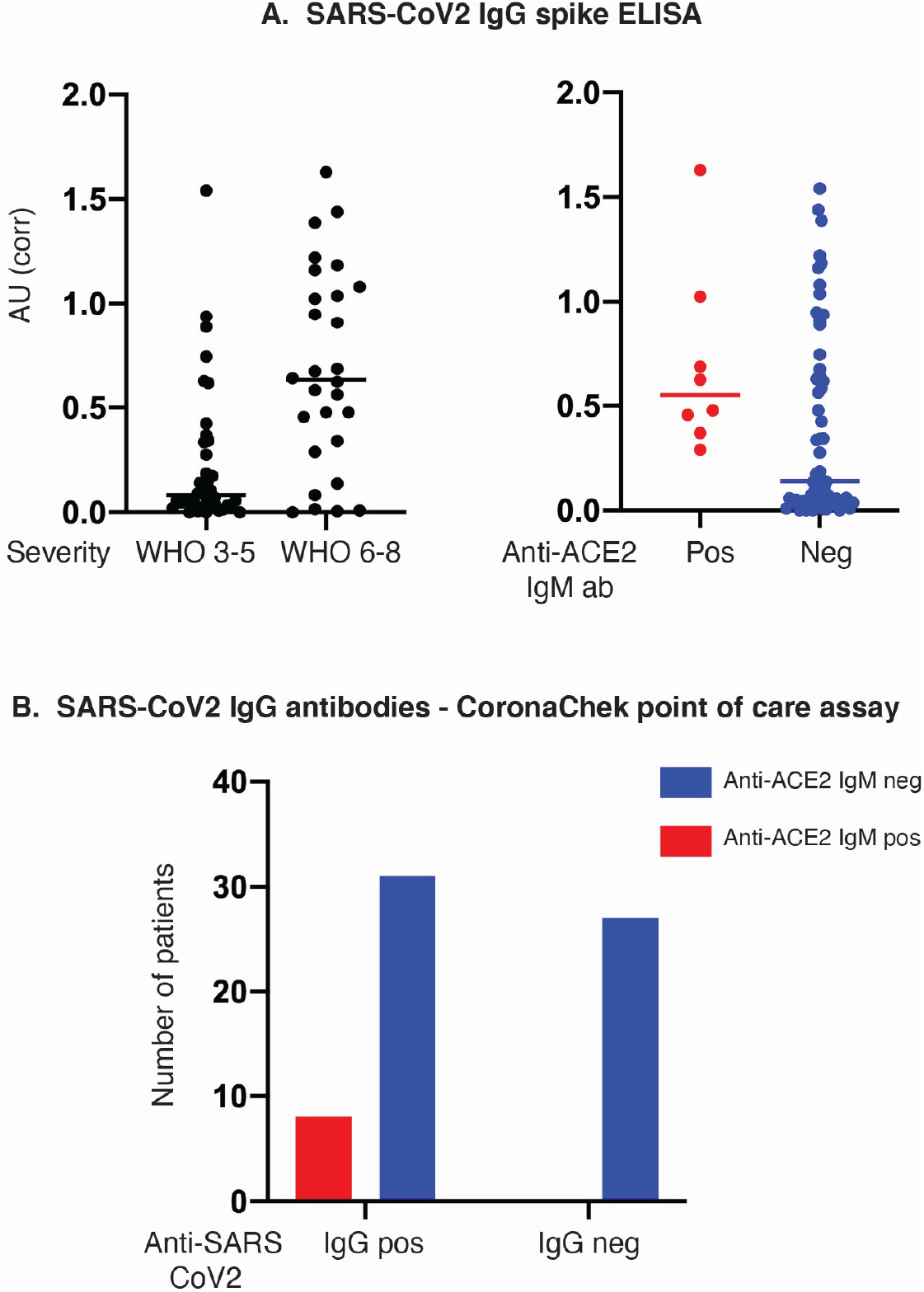
Antibodies against SARS-CoV-2 S-protein in anti-ACE2-positive COVID-19 patients. A: Anti-SARS-CoV-2 S-protein IgG antibodies were assayed by ELISA (N=66). Patients are shown grouped by disease severity in (left panel), and by anti-ACE2 IgM antibody status in (right panel). The mean ODs of anti-S antibodies were significantly higher in patients with severe compared to mild COVID (P<0.0001, Chi-squared). The median anti-S-antibody level was significantly higher in anti-ACE2 IgM-positive patients compared to anti-ACE2 IgM-negatives (P=0.028, Mann-Whitney test). B: Anti-S and -RBP antibodies assayed by the CoronaChek point of care assay. 8/8 (100%) of anti-ACE2 IgM-positive patients had a positive IgG result, compared to only 31/58 (53.4%) of anti-ACE2 IgM-negative patients (p=0.017, Fisher’s exact test). Red and blue denote anti-ACE2 IgM antibody-positive and -negative patients, respectively.

**Supplemental Fig 5:**
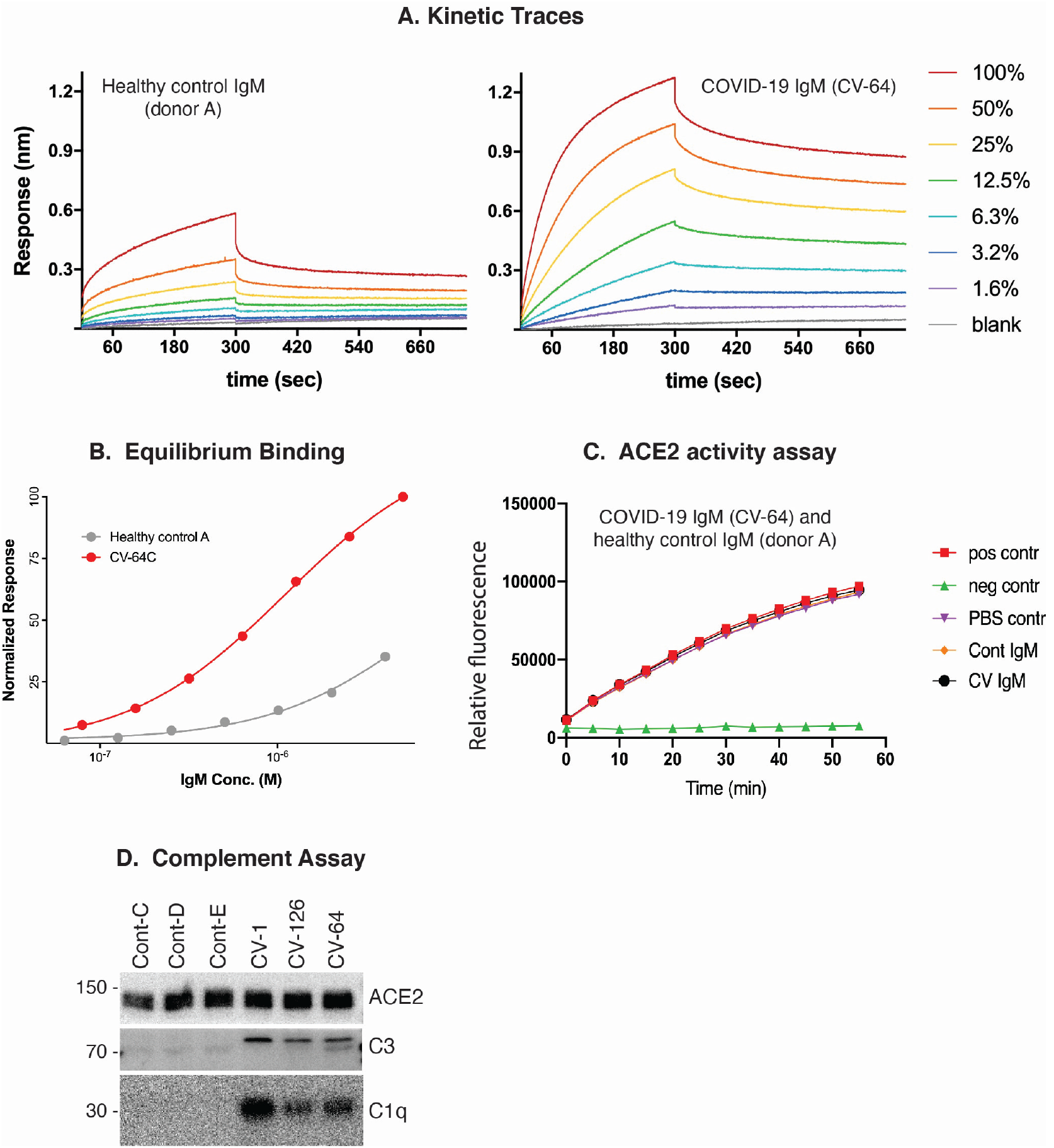
Properties of anti-ACE2 IgM antibodies. (A-B): Kinetics. A: Kinetic traces of the binding interactions between immobilized human ACE2 and purified IgM, as determined by biolayer interferometry. Percentages represent twofold dilutions of IgM from patient CV-64 and Control A. B: Equilibrium binding titrations. Normalized responses at the indicated concentrations of purified IgM from the donors shown in (A) are plotted (see Fig.3A&B for data obtained from CV-1 and control B). Kinetic parameters are provided in Fig. 3C. C: Anti-ACE2 IgM antibodies do not inhibit ACE2 activity. ACE2 activity, in the presence or absence of IgM from patient CV-64 or Control A, was measured using a fluorescent substrate in a time course assay. The positive control was ACE2 alone, and the negative control was ACE2 plus ACE2 inhibitor (see Fig 3C for data from CV-1 and control B). D: Complement activation induced by IgM antibodies to ACE2. Dynabeads containing immune complexes of ACE2 and purified IgM from controls (cont) or anti-ACE2 IgM from COVID-19 patients (CV) were incubated with human complement. Deposition of C1q and C3 was visualized by immunoblotting. ACE2 is shown as a loading control.

## Notes

### Competing Interest Statement

Antony Rosen and Livia Casciola-Rosen are listed as inventors on a patent application filed by Johns Hopkins University that encompasses aspects of this publication

### Author Declarations

These studies were approved by the JHU Institutional Review Board (IRB 00251725, IRB 00256018, 00256547), with a waiver of consent because all specimens and clinical data were de-identified by the Core for Clinical Research Data Acquisition of the Johns Hopkins Institute for Clinical and Translational Research; the study team had no access to identifiable patient data.

